# Trans-ethnic Polygenic Risk Scores for Body Mass Index: An International Hundred K+ Cohorts Consortium Study

**DOI:** 10.1101/2023.01.17.23284675

**Authors:** Huiqi Qu, John J Connolly, Peter Kraft, Jirong Long, Alexandre Pereira, Christopher Flatley, Constance Turman, Bram Prins, Frank Mentch, Paulo A Lotufo, Per Magnus, Meir J Stampfer, Rulla Tamimi, A Heather Eliassen, Wei Zheng, Gun Peggy Stromstad Knudsen, Oyvind Helgeland, Adam S. Butterworth, Hakon Hakonarson, Patrick M. Sleiman, the IHCC consortium

## Abstract

**Background:** While polygenic risk scores hold significant promise in estimating an individual’s risk of developing a complex trait such as obesity, their application in the clinic has, to date, been limited by a lack of data from non-European populations. As a collaboration model of the International Hundred K+ Cohorts Consortium (IHCC), we endeavored to develop a globally applicable trans-ethnic PRS for body mass index (BMI) through this relatively new international effort.

**Methods:** The PRS model was developed trained and tested at the Center for Applied Genomics (CAG) of The Children’s Hospital of Philadelphia (CHOP) based on a BMI meta-analysis from the GIANT consortium. The validated PRS models were subsequently disseminated to the participating sites. Scores were generated by each site locally on their cohorts and summary statistics returned to CAG for final analysis.

**Results:** We show that in the absence of a well powered trans-ethnic GWAS from which to derive SNPs and effect estimates, trans-ethnic scores can be generated from European ancestry GWAS using Bayesian approaches such as LDpred to adjust the summary statistics using trans-ethnic linkage disequilibrium reference panels. The ported trans-ethnic scores outperform population specific-PRS across all non-European ancestry populations investigated including East Asians and three-way admixed Brazilian cohort.

**Conclusions:** Widespread use of PRS in the clinic is hampered by a lack of genotyping data in individuals of non-European ancestry for the vast majority of traits. Here we show that for a truly polygenic trait such as BMI adjusting the summary statistics of a well powered European ancestry study using trans-ethnic LD reference results in a score that is predictive across a range of ancestries including East Asians and three-way admixed Brazilians.

## Introduction

Obesity is a global health issue^1^, with an adult prevalence of about 13% across the world (https://www.who.int/news-room/fact-sheets/detail/obesity-and-overweight). As a difficult condition to treat, obesity prevention is important which has led us to develop a trans-ethnic polygenic risk score (PRS) for body mass index (BMI) through the International HundredK+ Cohorts Consortium (IHCC)^2^. PRS aggregates the effects of many genetic variants across the human genome into a single score, which may effectively improve the prediction of a complex disease/trait and assist the differential diagnosis^3^. Our obesity PRS was based on the published GWAS meta-analysis of BMI that included 339,224 individuals of European ancestry. The study identified 97 genome-wide significant BMI-associated loci that account for approximately 2.7% of BMI variation alone. Genome-wide estimates suggest that common variation accounts for over 20% of variation in BMI. Various approaches for PRS calculation have been developed to date. The standard approaches for calculating risk scores involve linkage disequilibrium (LD)-based marker pruning followed by p-value thresholding of GWAS-based association statistics. While effective, these approaches lose information and can reduce predictive accuracy particularly where the test population differs in genetic ancestry from the GWAS sample. Bayesian approaches such LDpred^4^, a method that infers the posterior mean effect size of each marker by using a prior on effect sizes and LD information from an external reference panel may therefore improve prediction accuracy in multi-ethnic studies of diverse populations^4^. As most large scale GWAS have been conducted using only individuals of European ancestry there is a need to develop approaches that can port PRS using European ancestry derived effect estimates. More importantly, BMI and obesity are closely related to environmental factors^5^. It is essential to validate a multi-ethnic PRS in different regional populations, especially admixed populations. Leveraging an international effort supported by the IHCC that has brought together large scale cohorts with genotyping data from around the world, we explored the development of a LDpred-based, trans-ethnic (TE) obesity PRS through a collaboration of 6 international research centers (Table 1).

**Table 1.** Six international research centers for the development of the trans-ethnic PRS of BMI.

| **Cohort Name** | **PI - Lead** | **Participating countries** | **Current Enrollment** |
| --- | --- | --- | --- |
| **ELSA-Brasil** | Paulo A. Lotufo | Brazil: six cities | 15,105 |
| **Norwegian Mother and Child Cohort Study (MoBa)** | Per Magnus | Norway | 284,000 |
| **Children's Hospital of Philadelphia (CHOP) Biorepository** | Hakon Hakonarson | USA, Europe, South America, Canada, Saudi Arabia and Australia | 500,000 |
| **NHS (Nurses' Health Study, NCI)** | Meir Stampfer, Rulla Tamimi | USA | 121,700 |
| **NHSII (Nurses' Health Study II, NCI)** | Walter Willett, Heather Eliassen | USA | 116,430 |
| **Shanghai Men and Women's Health Study (2 cohorts)** | Wei Zheng | Shanghai, China | 136,000 |

## Methods

### Model training at the Children’s Hospital of Philadelphia (CHOP)

The PRS model development and training was carried out at the Center for Applied Genomics (CAG) according to the pipeline shown in Fig.1. SNP weights (i.e. posterior mean effect sizes) were calculated using Markov chain Monte Carlo (MCMC) Gibbs sampling^6^ as implemented in LDpred^4^. The summary statistics of genetic association with BMI were based on the meta-analysis of genome-wide association studies by the GIANT consortium^7^. We restricted the variants to SNPs included in the HapMap3 data. Five sets of SNP weights were generated based on the respective LD patterns from the following populations, African American (AA), Hispanic American (HAMR), East Asian (ASN), Northern European (EUR) and all of the above populations (i.e. trans-ethnic). For each group we selected 2500 CAG participants who clustered with the 1000 genome project reference data^8^ to generate the LD files. The trans-ethnic group included all populations tested. Each group of SNP weights includes 8 different sets corresponding respectively to the mixture probability values (i.e. fractions of causal markers) of [infinitesimal prior (LDpred-inf), 1, 0.3, 0.1, 0.03, 0.01, 0.003, and 0.001].

**Figure 1.**
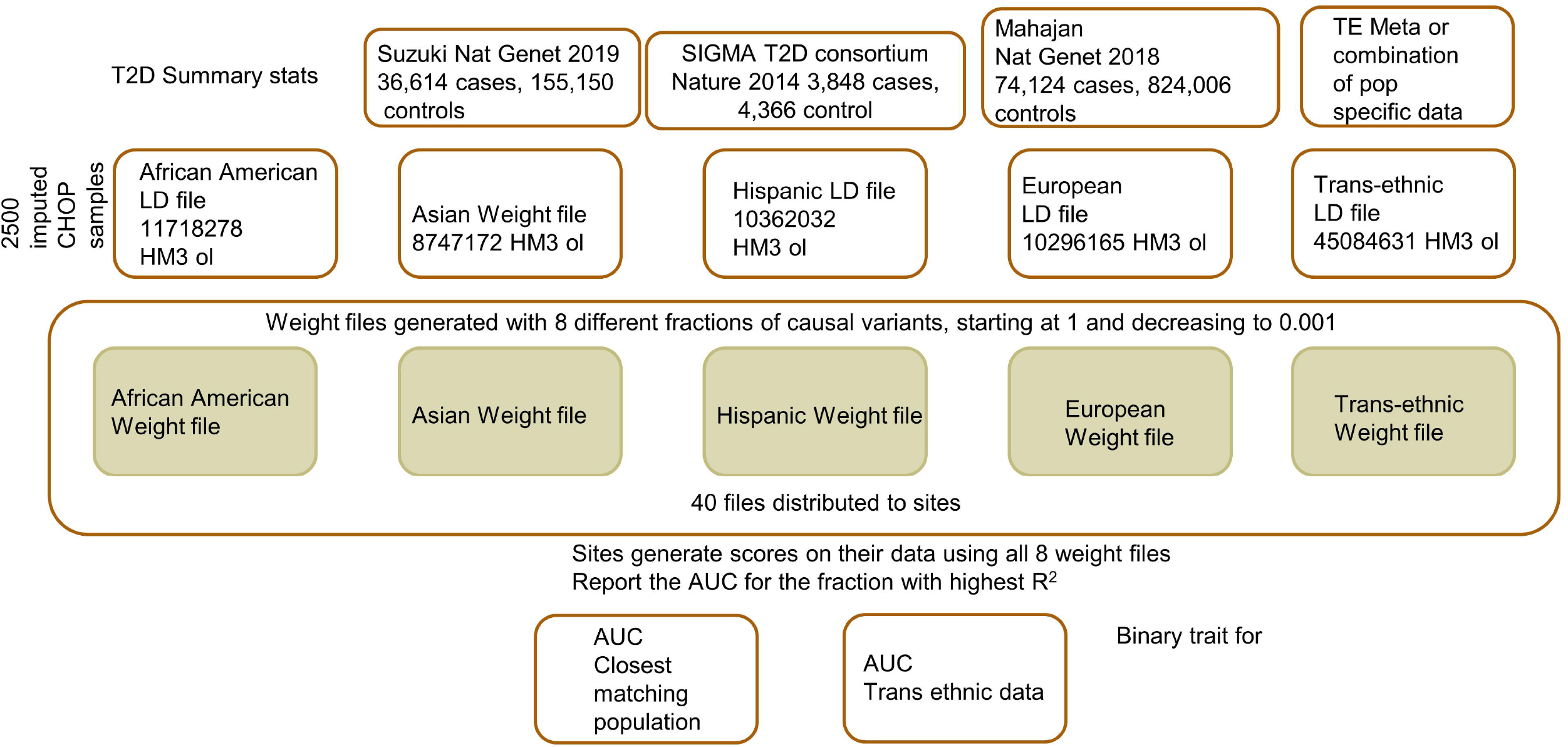
Development of a Transethnic Polygenic Risk Score for Body Mass Index.

### Initial validation of PRS models in-house

The initial assessment of the PRS models was based on the genomic data in-house at CAG, CHOP. The pediatric biobank built at CAG has archived samples from 500,000 children from USA, Europe, South America, Canada, Saudi Arabia and Australia^9^. For the initial validation of the trans-ethnic PRS model, 57,613 randomly selected individuals including individuals of European ancestry, African Americans, Hispanics/Latinos, and East / South Asians in order of frequency, with genotypes and BMI data from the CAG biobank were used for the validation. The principal component analysis (PCA) plot of the population structure is shown in Supplementary Figure 1. All the individuals have been genotyped with an Illumina Genotyping BeadChip with at least 550,000 SNPs genotyped. Genome-wide imputation was done using the TOPMed Imputation Server (https://imputation.biodatacatalyst.nhlbi.nih.gov) with the TOPMed (Version R2 on GRCh38) Reference Panel. Population ancestries of the research subjects were confirmed by principal component analysis (PCA) with genomic DNA markers, compared with the reference populations in the 1000 Genomes project^8^. Harmonization of SNP alleles in the PRS model was confirmed by comparing with the reference alleles of the TOPMed imputation.

### Validation of PRS models in regional populations

Having validated the trans-ethnic PRS models, we shared the protocol of the PRS models, as well as the SNPs and weights with the IHCC collaborators for assessment in 7 different cohorts of regional populations. The population sites included a three-way admixed Brazilian cohort from ELSA-Brasil^10 11^, the Norwegian Mother, Father and Child Cohort Study (MoBa, conducted by the Norwegian Institute of Public Health)^12^, two US based studies The Nurses’ Health Study (NHS)^13^ and Nurses’ Health Study II (NHSII)^14^, the UK based INTERVAL BioResource and two Chinese population samples from the Shanghai Men’s Health Study (SMHS) and Shanghai Women’s Health Study (SWHS)^15 16^(Table 1).

Each site generated the PRS on their cohort following the same protocol and using the same tools and the software packages. To compare the performance of trans-ethnic PRS vs. that of population-specific PRS in different regional populations, we requested each collaboration group run two calculations if possible, i.e., one PRS calculation for the specific population that is closest to their dataset, and one PRS for the trans-ethnic score. To do the dichotomous receiver operating characteristic (ROC) curve analysis, the top 5% and 1% of the BMI distribution within each study were defined as cases.

### Participating IHCC cohorts

The **Nurses’ Health Study I** recruited 121,700 married registered nurses in 1976. Blood samples were collected from 33,000 participants in 1989-90, and cheek cells from another 33,000 in 2001-4. Genome-wide association data are available on over 17,000 participants as part of studies of multiple complex diseases and traits, including breast cancer, type 2 diabetes, venous thromboembolism, and depression. All women included in these analyses are of European ancestry (cluster with European reference samples and do not self-report as other than European ancestry).

The **Nurses’ Health Study II** recruited 116,430 married registered nurses between the ages of 25 and 42 in 1989. Blood samples were collected from 29,000 participants in 1996-99, and cheek cells from another 30,000 in 2006. Genome-wide association data are available on over 12,000 participants as part of studies of multiple complex diseases and traits, including breast cancer, type 2 diabetes, venous thromboembolism, and depression. All women included in these analyses are of European ancestry (cluster with European reference samples and do not self-report as other than European ancestry).

Body mass index was self-reported at time of blood draw or cheek cell collection. Diabetes cases were defined as self-reported diabetes confirmed by a validated supplementary questionnaire.

The **Shanghai Women’s Health Study** (**SWHS**) is a large population-based prospective cohort study initiated in 1996 ^15^. Approximately 75,000 Chinese women who lived in Shanghai were recruited into the study. In addition to survey data, blood and urine samples were collected from most study participants at the baseline recruitment.

The **Shanghai Men’s Health Study** (**SMHS**) is a population-based cohort study of 61,480 Chinese men between ages 40 and 74 who lived in eight urban communities in Shanghai at enrollment (2002-2006) ^17^. Detailed information on dietary and other lifestyle factors was collected at baseline and updated in follow-up surveys. Biological samples (blood, and or urine) were collected from 89% of cohort members.

The **Norwegian Mother, Father and Child Cohort Study (MoBA)** was established and is conducted by the Norwegian Institute of Public Health (NIPH). MoBa is an ambitious family-oriented cohort study that aims to find causes of diseases and explain trajectories and variability of health-related traits over a life-course span. Between 1999 - 2008 pregnant women were invited to take part in the study around the time of the ultrasound examination in week 17-20 of gestation. The fathers of the children were also invited to participate. Biological material has been collected from mothers, fathers and children and has been stored in a biobank. Self-reported data are collected from regular questionnaires about general health, diet and environmental exposure. The cohort includes approximately 109,000 children, 91,000 women and 71,700 men. 50,290 Northern European adult males and females were analysed in this study, with mean BMI=24.96(SD=3.9).

### The Brazilian Longitudinal Study of Adult Health (ELSA-Brasil)

enrolled 15,105 civil servants aged 35 to 74 years-old living in six cities ^10^, addressing the incidence of non-communicable diseases. From the 15105 participants, 9333 DNA samples were analyzed for genetic ancestry using a software tool for maximum likelihood estimation of individual ancestries from multilocus SNP genotype datasets^11^.

The **INTERVAL BioResource** recruited 45,263 whole blood donors (22,466 men and 22,797 women) between June 11, 2012, and June 15, 2014^18^. Donors were aged 18 years or older from 25 NHS Blood and Transplant (NSHBT) blood donation centres distributed across England, UK. Donors provided blood samples at baseline to enable DNA extraction and self-reported their height and weight for estimation of BMI. Genotyping was conducted using the Affymetrix Axiom UK Biobank array with imputation using a combined 1000 Genomes Phase 3 / UK10K reference panel. A total of 38,319 European ancestry adult participants were included in the final analyses

## Results

### Initial validation of PRS models in-house

The performance of the trans-ethnic PRS model tested in-house is shown in Fig.2. As shown by our analysis, AUC>0.720 to predict top 1% BMI was achieved in all the PRS models with mixture probability≥0.03. The highest AUC is seen at the mixture probabilities of 0.03 to 0.1.

**Figure 2.**
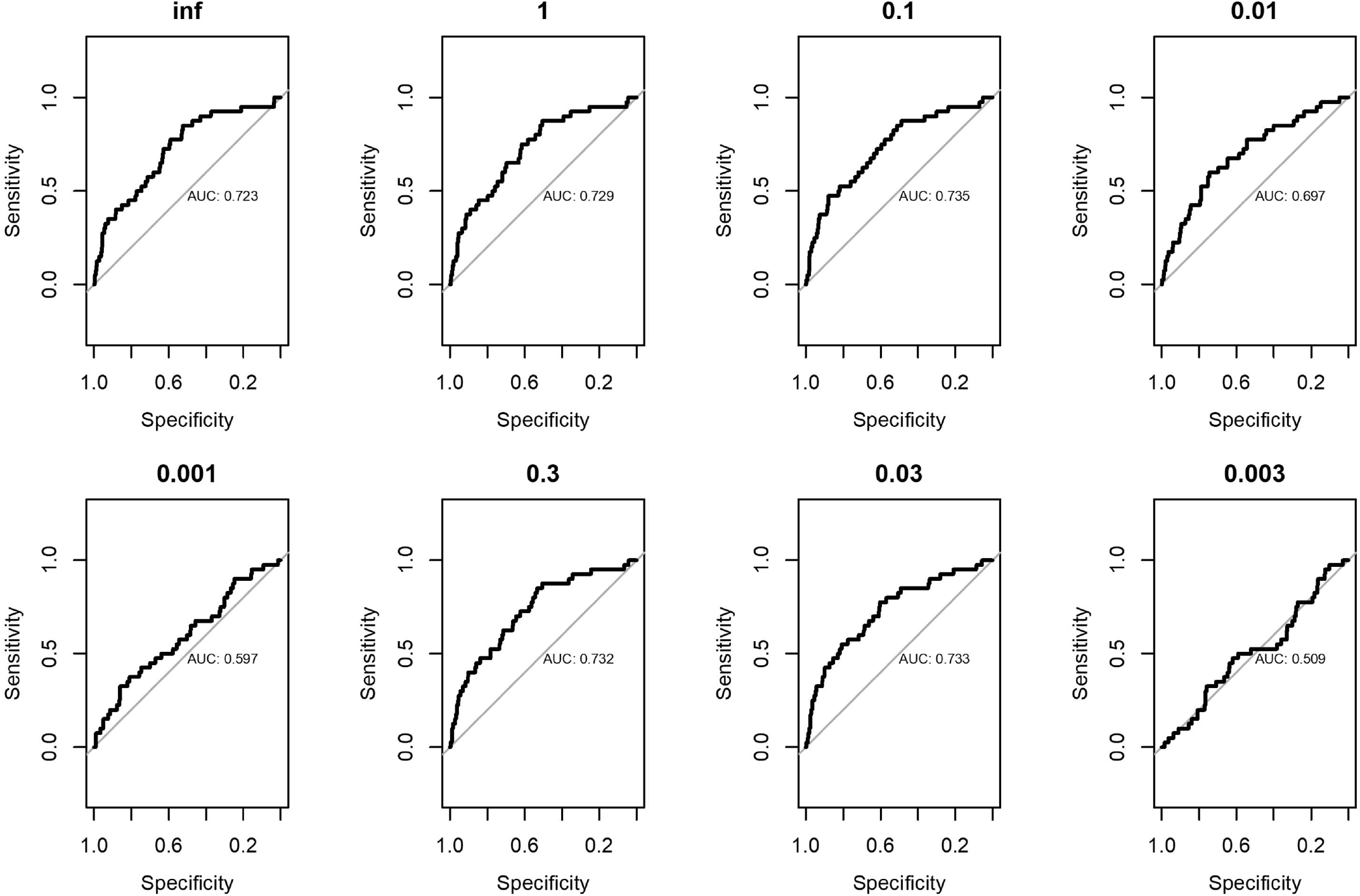
AUC values following the initial validation of PRS models in-house. 57,613 randomly selected individuals with genotypes and BMI data from the the Center for Applied Genomics biobank at the Children’s Hospital of Philadelphia were used for the validation.

### Validation of the trans-ethnic PRS models in regional populations

By the Chinese population in the SMHS and SWHS cohorts, the PRS models were tested in 866 men in the SMHS cohort and 3,120 women in the SWHS cohort and the combination of both. Across all three analyses the trans-ethnic (TE) score outperformed the ancestry specific East-Asian reference with a TE AUC of 0.76 compared with 0.479 for the ASN reference in men, TE AUC of 0.737 vs ASN AUC of 0.719 in women, and a TE AUC of 0.717 vs ASN AUC of 0.706 in the combined dataset at the most informative fraction (Table 2).

**Table 2.** BMI PRS test by the Shanghai Men and Women’s Health Study.

| Cohorts | Racial groups of weights | SMHS (N=866) |  |  | SWHS (N=3,120) |  |  | SWMHS (N=3,986) |  |  |
| --- | --- | --- | --- | --- | --- | --- | --- | --- | --- | --- |
|  |  | P* | AUC | P # | P* | AUC | P # | P* | AUC | P # |
| **INF** | ASN | 0.07 | 0.667 | 1.12E-10 | 6.61E-06 | 0.725 | 5.21E-33 | 1.91E-06 | 0.706 | 4.66E-42 |
|  | TE | 0.044861 | 0.684 | 3.03E-11 | 4.88E-06 | 0.732 | 1.80E-32 | 1.15E-06 | 0.717 | 5.41E-42 |
| **1** | ASN | 0.05 | 0.682 | 1.99E-11 | 1.71E-05 | 0.714 | 3.92E-33 | 7.66E-06 | 0.694 | 7.74E-43 |
|  | TE | 0.040165 | 0.684 | 1.91E-11 | 5.09E-06 | 0.728 | 1.32E-33 | 1.78E-06 | 0.709 | 2.43E-43 |
| **0.3** | ASN | 0.03 | 0.707 | 3.40E-12 | 1.10E-05 | 0.719 | 2.85E-34 | 4.90E-06 | 0.699 | 1.13E-44 |
|  | TE | 0.030129 | 0.692 | 4.93E-12 | 2.54E-06 | 0.737 | 1.65E-35 | 1.16E-06 | 0.713 | 8.18E-46 |
| **0.1** | ASN | 0.02 | 0.721 | 2.10E-13 | 8.01E-06 | 0.722 | 1.83E-35 | 3.55E-06 | 0.702 | 6.26E-47 |
|  | TE | 0.017299 | 0.716 | 2.96E-13 | 2.15E-06 | 0.735 | 8.27E-37 | 1.21E-06 | 0.711 | 3.32E-48 |
| **0.03** | ASN | 0.03 | 0.719 | 8.04E-15 | 3.62E-05 | 0.703 | 2.24E-35 | 1.94E-05 | 0.684 | 5.40E-48 |
|  | TE | 0.015892 | 0.733 | 1.10E-14 | 1.34E-05 | 0.714 | 3.65E-37 | 8.83E-06 | 0.694 | 9.08E-50 |
| **0.01** | ASN | 0.22 | 0.65 | 0.04 | 0.98 | 0.515 | 0.08 | 0.82 | 0.497 | 0.01 |
|  | TE | 0.11957 | 0.647 | 0.96 | 0.57 | 0.533 | 0.15 | 0.88 | 0.516 | 0.19 |
| **0.003** | ASN | 0.21 | 0.622 | 0.41 | 0.91 | 0.517 | 0.36 | 0.35 | 0.471 | 0.64 |
|  | TE | 0.73 | 0.527 | 0.28 | 0.66 | 0.541 | 0.15 | 0.35 | 0.561 | 0.08 |
| **0.001** | ASN | 0.87 | 0.479 | 0.91 | 0.27 | 0.568 | 0.2 | 0.29 | 0.569 | 0.22 |
|  | TE | 0.017789 | 0.76 | 0.13 | 0.66 | 0.534 | 0.23 | 0.42 | 0.551 | 0.08 |

\* Logistic regression analyses were performed after coding top 99% distribution of BMI as cases and the remainings as controls. # Linear regression analyses were performed using BMI as continuous variable.

The Brazilian population in the ELSA-Brasil cohort is three way admixed. We therefore compared the trans-ethnic score in this population to each of the three founders: American, African and European. The trans-ethnic score outperformed all three founder population ancestry-specific scores at the most informative fraction of 0.03 at both 95^th^ and 99^th^ percentiles (Table 3).

**Table 3.**
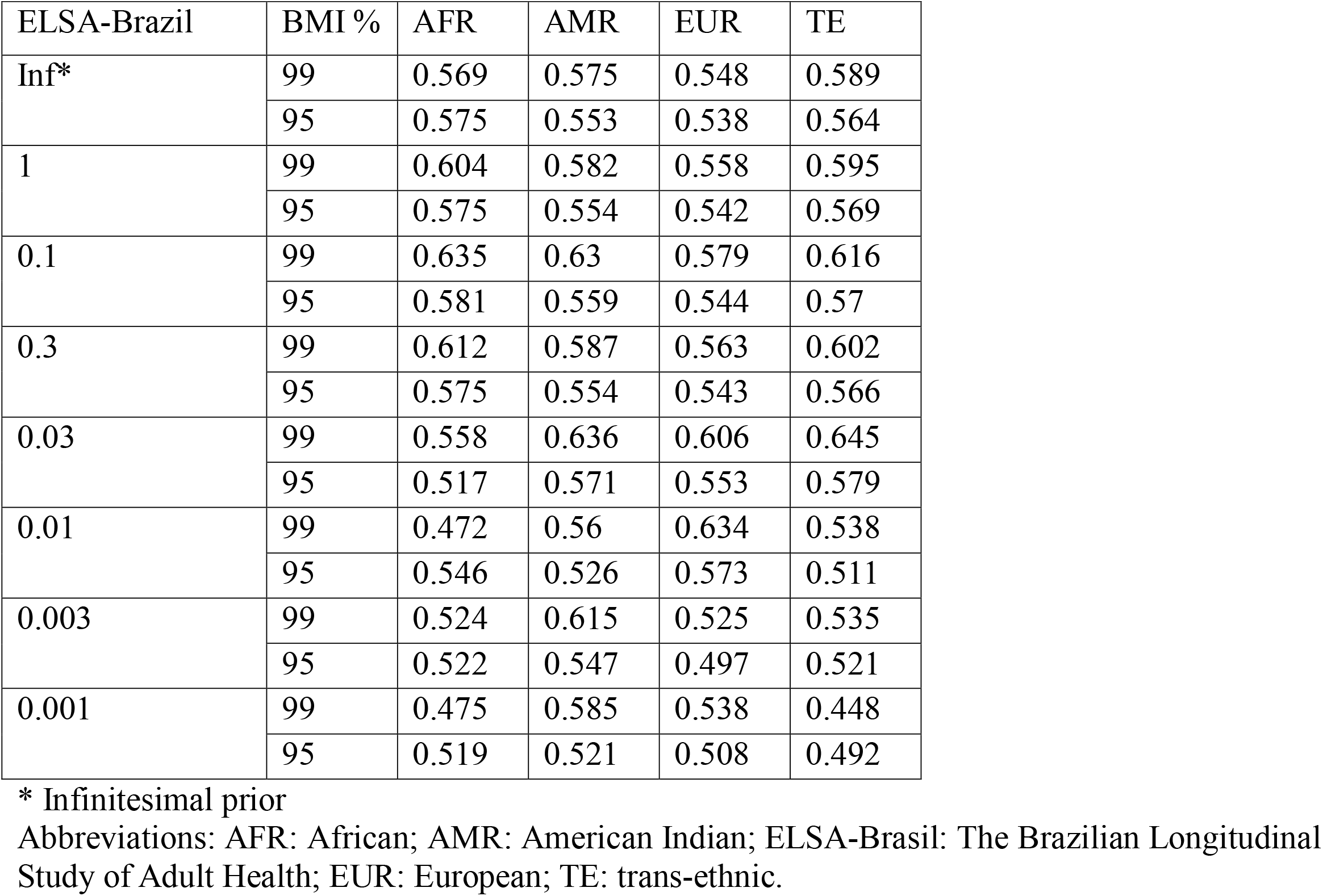
BMI PRS test by the Brazilian population in the ELSA-Brasil cohort.

| ELSA-Brazil | BMI % | AFR | AMR | EUR | TE |
| --- | --- | --- | --- | --- | --- |
| Inf* | 99 | 0.569 | 0.575 | 0.548 | 0.589 |
|  | 95 | 0.575 | 0.553 | 0.538 | 0.564 |
| 1 | 99 | 0.604 | 0.582 | 0.558 | 0.595 |
|  | 95 | 0.575 | 0.554 | 0.542 | 0.569 |
| 0.1 | 99 | 0.635 | 0.63 | 0.579 | 0.616 |
|  | 95 | 0.581 | 0.559 | 0.544 | 0.57 |
| 0.3 | 99 | 0.612 | 0.587 | 0.563 | 0.602 |
|  | 95 | 0.575 | 0.554 | 0.543 | 0.566 |
| 0.03 | 99 | 0.558 | 0.636 | 0.606 | 0.645 |
|  | 95 | 0.517 | 0.571 | 0.553 | 0.579 |
| 0.01 | 99 | 0.472 | 0.56 | 0.634 | 0.538 |
|  | 95 | 0.546 | 0.526 | 0.573 | 0.511 |
| 0.003 | 99 | 0.524 | 0.615 | 0.525 | 0.535 |
|  | 95 | 0.522 | 0.547 | 0.497 | 0.521 |
| 0.001 | 99 | 0.475 | 0.585 | 0.538 | 0.448 |
|  | 95 | 0.519 | 0.521 | 0.508 | 0.492 |

\* Infinitesimal prior
Abbreviations: AFR: African; AMR: American Indian; ELSA-Brasil: The Brazilian Longitudinal Study of Adult Health; EUR: European; TE: trans-ethnic.

### Evaluation of the PRS in the four European ancestry cohorts

Across all four European ancestry cohorts both European-specific PRS models and the trans-ethnic PRS models had the largest AUCs with a mixture probability of 0.03. Across the three cohorts where both the European-specific PRS model and the trans-ethnic PRS model were run, (INTERVAL, NHS and NHSII), the performance of the scores was comparable (Table 4).

**Table 4.** BMI PRS test by the NHS and NHSII cohorts*.

| **top 1%/P-0.03.AUC** | **EUR-NHS1** | **TE-NHS1** |
| --- | --- | --- |
| **NHS** | 0.730 | 0.727 |
| **NHSII** | 0.736 | 0.735 |
| **top 5%/P-0.03.AUC** |  |  |
| **NHS** | 0.703 | 0.701 |
| **NHSII** | 0.721 | 0.718 |

\* NHS: the Nurses' Health Study.

Only the European-specific PRS models were tested in the Norwegian MoBa study. The PRS model showed AUC≥0.686 to predict top 5% BMI, and AUC≥0.735 to predict top 1% BMI, in the models with mixture probability≥0.03 (Table 5).

**Table 5.** BMI PRS test by the Norwegian Mother and Child Cohort Study (MoBA)

| **MoBa** | **95%**<br>**BMI** | **99%**<br>**BMI** |
| --- | --- | --- |
| **Inf\*** | 0.686 | 0.735 |
| **1** | 0.689 | 0.738 |
| **0.3** | 0.694 | 0.745 |
| **0.1** | 0.699 | 0.751 |
| **0.03** | 0.704 | 0.756 |
| **0.01** | 0.658 | 0.7 |
| **0.003** | 0.594 | 0.626 |
| **0.001** | 0.552 | 0.565 |

\* Infinitesimal prior

## Discussion

In order for polygenic risk scores to achieve their clinical potential, and avoid exacerbating health disparities due to the lack of genomic information in minorities^19^, they have to be universally applicable regardless of a patient’s genetic ancestry. Ideally, trans-ethnic PRS would be generated from true trans-ethnic GWAS however, well powered trans-ethnic studies remain the exception for the majority of traits and phenotypes. In this study we evaluated the performance of a BMI PRS that is based on European ancestry GWAS effect sizes combined with trans-ethnic LD patterns using a Bayesian approach as implemented in LDpred^4^. In a federated model, we developed the PRS score at CAG and disseminated the standard operating procedure along with the SNPs and weights files and the population specific LD matrices to participant sites within the IHCC. This model allowed us to quickly test the hypothesis in various world populations without the need for data transfer and hence time-consuming data sharing agreements. By providing detailed protocols and all required files to run the scores, we made efforts to minimize the work load on each collaboration group while enabling direct comparisons between groups.

It is worth to mention that, in contrast to the LDpred^4^ used in this study, a new version of the method LDpred2 has been released^20^. LDpred2 made a significant effort to address the potential bias of Gibbs sampling in the human leukocyte antigen (*HLA*) region at chromosome 6 with extended LD^20^. In contrast, the *HLA* region has been removed in the LDpred modeling. The *HLA* region is highly polymorphic, with highly diverse frequencies across different populations, as well as extended and strong LD due to significant evolutionary selection pressure in human populations^21 22^. Including the *HLA* region will cause significant difference of PRS across different ethnicities, as we have observed in the trans-ethnic scoring of an autoimmune disease, type 1 diabetes (T1D), with *HLA* as a major risk factor^23^. In addition, there has been no GWAS study to date suggesting the role of the *HLA* region in BMI or obesity. Nevertheless, it is interesting to leverage the IHCC resources to examine the performance of LDpred2 in the trans-ethnic scoring of BMI.

As the results show, our trans-ethnic PRS models outperformed the ancestry specific models in the non-European populations tested including Chinese and Brazilian. Importantly the UK and US data from the INTERVAL and NHS cohorts demonstrate that there is no appreciable loss in predictive power in European ancestry individuals when using a trans-ethnic score. As such, and in the absence of trans-ethnic effect sizes from diverse GWAS, we propose that using trans-ethnic reference data to adjust the summary statistics for the effects of LD patterns improves the performance of PRS in populations that have not been included in the generation of the summary stats. On the other hand, in the absence of reference panels for different populations, the genome-wide LD scores and matrices from the Pan-UK Biobank resource^24^, or calculated by genome sequencing data of the Genome Aggregation Database (gnomAD)^25^, may provide sufficient resolution.

As the first effort by the IHCC to leverage the existing datasets that reside within this large scale consortium for a transethnic PRS on BMI, we envision an opportunity to scale this to other cohorts within the consortium and expand the number of traits that can be analyzed. As such, the IHCC presents a rich resource of data for collaborative research with transethnic focus, where there is much unmet need at the present time and an area of research that has been largely ignored.

## Data Availability

Supporting data from this study can be obtained by emailing the corresponding author Dr. Hakon Hakonarson.

## Acknowledgements

We thank all the participants who contributed to and enabled this study.

## Ethical Approval

This study was exempted by the Institutional Review Board (IRB) of the Children’s Hospital of Philadelphia. Human participants and personal information are inaccessible to the research group due to sample and data deidentification. All human subjects or their proxies provided written informed consent for the respective studies involved.

## Consent for publication

All authors have provided consent for publication of the manuscript.

## Figure Legend

**Supplementary Figure 1**. The principal component analysis (PCA) plot of the population structure of CAG samples for initial validation.

## Reference

1. Caballero B. The global epidemic of obesity: an overview. Epidemiologic reviews 2007;29(1):1–5.

2. Manolio TA, Goodhand P, Ginsburg G. The International Hundred Thousand Plus Cohort Consortium: integrating large-scale cohorts to address global scientific challenges. The Lancet Digital health 2020;2(11):e567–e68. doi: 10.1016/s2589-7500(20)30242-9 [published Online First: 2020/10/27]

3. Lambert SA, Abraham G, Inouye M. Towards clinical utility of polygenic risk scores. Human Molecular Genetics 2019;28(R2):R133–R42. doi: 10.1093/hmg/ddz187

4. Vilhjálmsson BJ, Yang J, Finucane HK, et al. Modeling linkage disequilibrium increases accuracy of polygenic risk scores. The American Journal of Human Genetics 2015;97(4):576–92.

5. Barness LA, Opitz JM, Gilbert-Barness E. Obesity: genetic, molecular, and environmental aspects. American journal of medical genetics part A 2007;143(24):3016–34.

6. Carlo CM. Markov chain monte carlo and gibbs sampling. Lecture notes for EEB 2004;581:540.

7. Locke AE, Kahali B, Berndt SI, et al. Genetic studies of body mass index yield new insights for obesity biology. Nature 2015;518(7538):197–206. doi: 10.1038/nature14177 [published Online First: 2015/02/13]

8. Siva N. 1000 Genomes project. Nature biotechnology 2008;26(3):256–57.

9. Connolly JJ, Glessner JT, Li D, et al. The Center for Applied Genomics at The Children’s Hospital of Philadelphia–Pediatric Perspectives on Genomic Medicine adminpmls2016 2021-03-04T16: 43: 43-05: 00 The Center for Applied Genomics at The Children’s Hospital of Philadelphia–Pediatric Perspectives on Genomic Medicine. 2020

10. Aquino EM, Barreto SM, Bensenor IM, et al. Brazilian longitudinal study of adult health (ELSA-Brasil): objectives and design. American journal of epidemiology 2012;175(4):315–24.

11. Chor D, Pereira A, Pacheco AG, et al. Context-dependence of race self-classification: Results from a highly mixed and unequal middle-income country. PLOS ONE 2019;14(5):e0216653. doi: 10.1371/journal.pone.0216653

12. Magnus P, Birke C, Vejrup K, et al. Cohort profile update: the Norwegian mother and child cohort study (MoBa). International journal of epidemiology 2016;45(2):382–88.

13. Colditz GA, Hankinson SE. The Nurses’ Health Study: lifestyle and health among women. Nature reviews Cancer 2005;5(5):388–96. doi: 10.1038/nrc1608 [published Online First: 2005/05/03]

14. Eliassen AH, Spiegelman D, Hollis BW, et al. Plasma 25-hydroxyvitamin D and risk of breast cancer in the Nurses’ Health Study II. Breast Cancer Research 2011;13(3):1–7.

15. Zheng W, Chow W-H, Yang G, et al. The Shanghai Women’s Health Study: rationale, study design, and baseline characteristics. American journal of epidemiology 2005;162(11):1123–31.

16. Shu XO, Li H, Yang G, et al. Cohort Profile: The Shanghai Men’s Health Study. Int J Epidemiol 2015;44(3):810–8. doi: 10.1093/ije/dyv013 [published Online First: 2015/03/04]

17. Shu X-O, Li H, Yang G, et al. Cohort profile: the Shanghai men’s health study. International journal of epidemiology 2015;44(3):810–18.

18. Di Angelantonio E, Thompson SG, Kaptoge S, et al. Efficiency and safety of varying the frequency of whole blood donation (INTERVAL): a randomised trial of 45□000 donors. Lancet (London, England) 2017;390(10110):2360–71. doi: 10.1016/s0140-6736(17)31928-1 [published Online First: 2017/09/25]

19. Martin AR, Kanai M, Kamatani Y, et al. Clinical use of current polygenic risk scores may exacerbate health disparities. Nature genetics 2019;51(4):584–91. doi: 10.1038/s41588-019-0379-x [published Online First: 2019/03/29]

20. Privé F, Arbel J, Vilhjálmsson BJ. LDpred2: better, faster, stronger. Bioinformatics (Oxford, England) 2020;36(22-23):5424–31. doi: 10.1093/bioinformatics/btaa1029 [published Online First: 2020/12/17]

21. Prugnolle F, Manica A, Charpentier M, et al. Pathogen-driven selection and worldwide HLA class I diversity. Current biology 2005;15(11):1022–27.

22. Meyer D, Vr CA, Bitarello BD, et al. A genomic perspective on HLA evolution. Immunogenetics 2018;70(1):5–27. doi: 10.1007/s00251-017-1017-3 [published Online First: 2017/07/09]

23. Qu HQ, Qu J, Glessner J, et al. Improved genetic risk scoring algorithm for type 1 diabetes prediction. Pediatric Diabetes 2022;23(3):320–23.

24. Pan-UKB team. https://pan.ukbb.broadinstitute.org. 2020

25. Karczewski K, Francioli L. The genome aggregation database (gnomAD). MacArthur Lab 2017:1–10.

