## Supplementary figures and images for "Trans-ethnic Polygenic Risk Scores for Body Mass Index: An International Hundred K+ Cohorts Consortium Study"

### Supplementary Figure 1

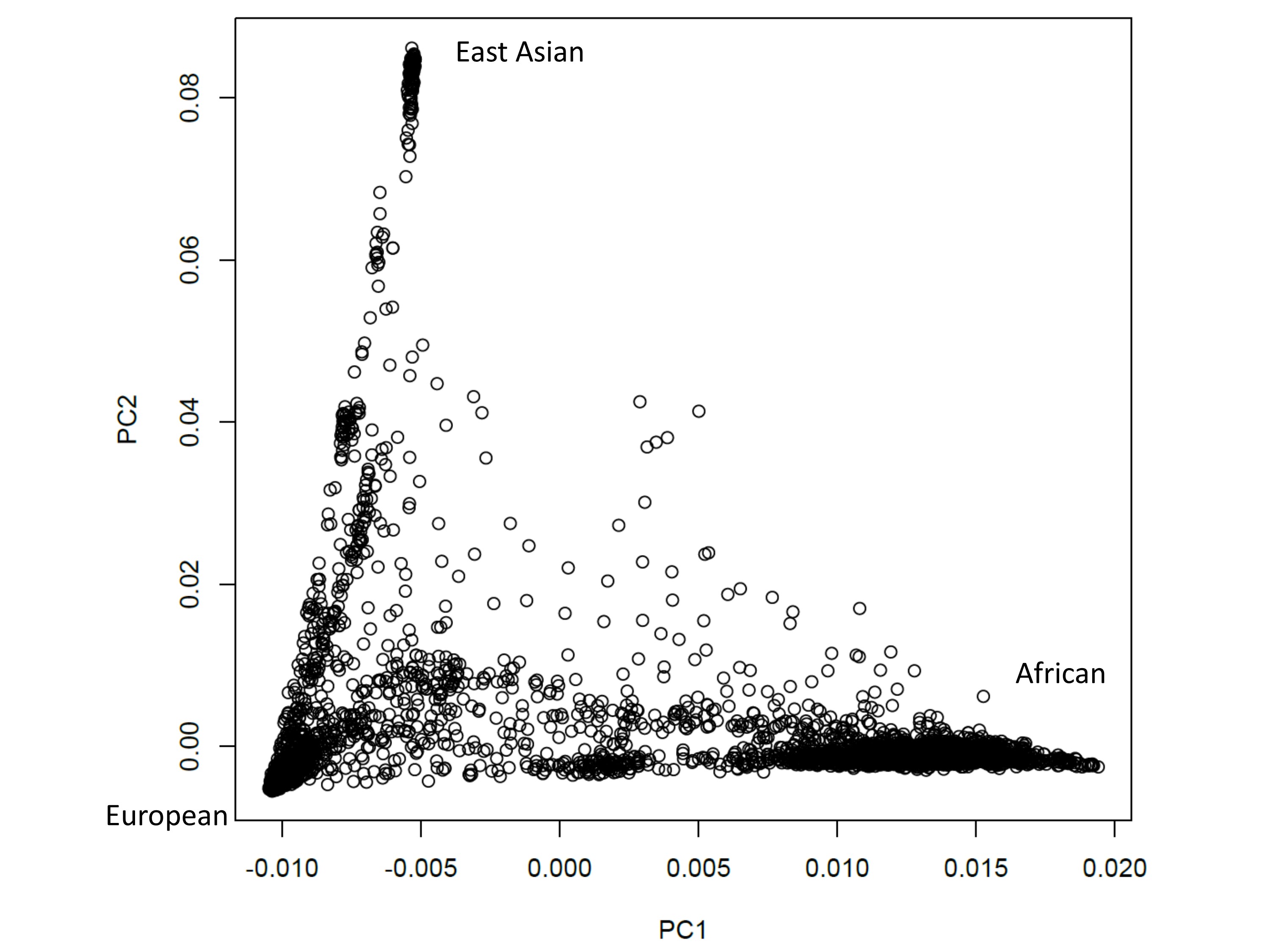
